# Follow-Up In Patients With Respiratory Disability After ARDS Related To COVID-19: A Systematic Review

**DOI:** 10.1101/2023.06.24.23291855

**Authors:** Panagiota Triantafyllaki, Marios Charalampopoulos, Christina-Athanasia Sampani, Christos Triantafyllou, Dimitrios Papageorgiou

**Affiliations:** General Hospital of Athens “Evangelismos”, Athens, Greece; Medecins Sans Frontieres – Greece; General Hospital of Athens “G. Gennimatas”, Athens, Greece; National and Kapodistrian University of Athens, School of Health Science, Department of Nursing, Goudi Athens, Greece; School of Health and Care Sciences, Department of Nursing, University of West Attica

**Keywords:** ARDS, SARS-CoV-2, ICU, COVID-19, follow up, respiratory function

## Abstract

**Introduction:** Acute Respiratory Distress Syndrome (ARDS) is an acute inflammatory pulmonary process that leads to protein-rich, non-hydrostatic pulmonary edema, undesirable hypoxemia, and lung stiffness. Due to COVID-19, a significant proportion of people who will require hospitalization to treat COVID-19, between 15%-30%, will develop severe respiratory failure, ARDS, and an increased likelihood of intubation for mechanical respiratory support.

**Aim:** To investigate the pulmonary function in COVID-19-related ARDS survivors after hospitalization.

**Methods:** A search was performed on the Greek and international literature, as well as at the online Databases PubMed, Cochrane, Embase, and Google Scholar. Exclusion and integration criteria were set for the studies found, and a flow chart was created for the studies included.

**Results:** Through the search, 352 articles were found matching the subject under study, and after further evaluation, four articles were included. The majority of the articles highlight that after ARDS occurs due to COVID-19, patients face impaired pulmonary function in combination with other physical and psychological symptoms like weakness, anxiety, depression, and generalized functional disability.

**Conclusions:** It is a fact that COVID-19 disease, in severe form and following the need for hospitalization due to the development of ARDS, results in an increased likelihood of prolonged occurrence of some symptoms of impaired respiratory function. Impaired CO2 diffusion is observed in the majority of studies as well as impaired respiratory function regarding prolonged imaging findings and impaired physical function.

## Introduction

Acute Respiratory Distress Syndrome (ARDS) is an acute inflammatory pulmonary process that leads to protein-rich, non-hydrostatic pulmonary edema, undesirable hypoxemia, and lung stiffness. As a result of the above, there is an inability to eliminate carbon dioxide and a disturbance in gas exchange. According to the epidemiological data on the disease, there is a dramatic variability in the incidence of the disease between different continents. In particular, in South America, the incidence of the disease is 10,1/100 000 person-years, in Europe 17,9/100 000 person-years, in Australia 34/100 000 person-years and in the USA 78,9/100 000 person-years, with relative geographical variability. However, a consistent difference occurs in European countries, where in Finland, the incidence of the disease is recorded at 10.6/100,000 person-years, while in Sweden at 17.9/100,000 person-years and in Spain at 25.5/100,000 person-years. These rates are derived from surveys based on hospitalization of patients in whom ARDS incidence ranged from 7.1% to 12.5% of the total incidence of all ICU cases in Europe.^1^

ARDS is a syndrome with multiple risk factors, resulting in acute respiratory failure. Most cohort studies suggest a pattern of clinical risk factors, with pneumonia being the major risk factor (35-50%), followed by pulmonary sepsis (30%), aspiration (10%), and trauma (10%). In addition to the above clinical risk factors, age (> 43 years), racial identity (non-white race), and genetic background are associated with higher rates of the disease.^2^

Clinically, it is characterized by acute respiratory failure, bilateral infiltrates on chest X-ray, and hypoxemia with an evaluation of PaO2/FiO2mmHg. However, there shall be no evidence of left atrial hypertension or pulmonary capillary hypertension.^3^

A new pathogen responsible for severe ARDS is the new SARS-CoV-2 coronavirus, which is genetically identical to SARS-CoV to a significant extent. The new coronavirus emerged in China in early 2020, and since then, there has been a significant spread, eventually causing a pandemic. As a virus mainly affecting the respiratory system, it manifests with fever, headache, dry cough, generalized weakness, and fatigue symptoms. A significant proportion of people who will require hospitalization to treat Covid-19, between 15%-30%, will develop severe respiratory failure, ARDS, and an increased likelihood of intubation for mechanical respiratory support,^4,5,6^

## Methods

We collected data from the international electronic scientific bases PubMed, SCOPUS, and EMBASE as well as at the Greek IATROTEK0online, both in English and Greek languages, respectively. No limitations were applied regarding the articles’ publication time. International or local locations of survey appliances were included. The Keywords used were «ARDS, SARS-CoV-2, ICU, COVID-19, follow up, pulmonary function».

## Results

In the absence of Greek literature, this article is based mainly on English studies without time limitations. The articles met the inclusion criteria and were inserted into this systematic review 4. The above results and the flow chart of the studies included are analyzed below (Figure 1).

**Figure 1:**
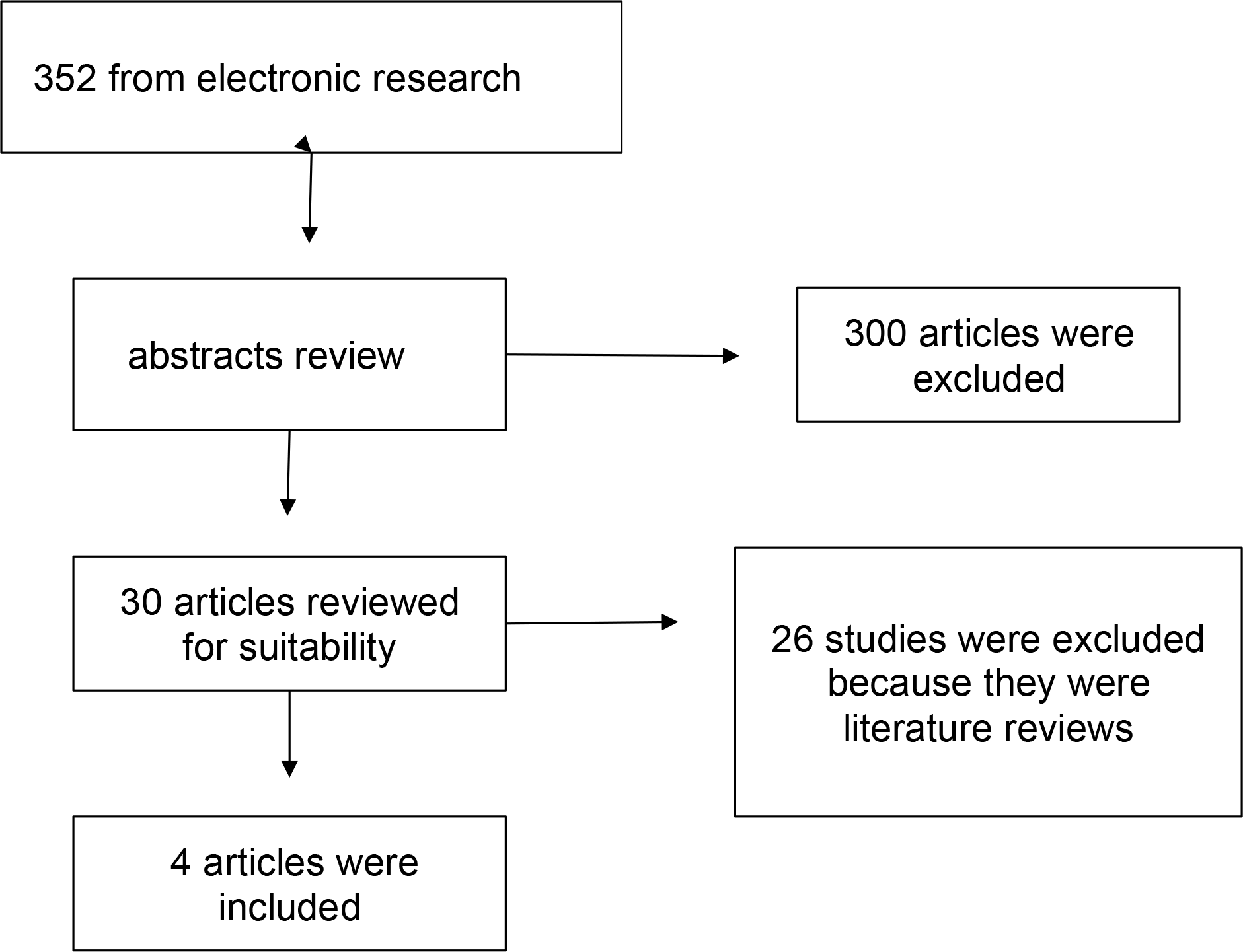
Flow chart of the systematic review.

### R Torres-Castro et al. (2020)^7^

In a systematic review conducted on long-term respiratory functionality in patients hospitalized for Covid 19, seven (7) studies and a total of 380 patients were included. In these patients, respiratory function assessment tests using a spirometer were performed between one (1) week and three (3) months after discharge. The respiratory parameters calculated were dynamic, vital capacity (FVC), the fast expiratory volume of air in one minute (FEV1), FEV1/FVC ratio, total lung capacity (TLC), residual capacity (RV), vital capacity (VC) and diffusing capacity. In the results, in 40% of subjects, impaired lung diffusing capacity was observed. In 15%, respiratory function limitation was observed, and in 7%, respiratory function was consistent with obstructive disease.

### Yu-Miao Zhao et al. (2020)^8^

In a different retrospective multicenter cohort study in China, respiratory function in surviving COVID-19 patients was studied over a three-month period. A total of 55 patients were enrolled, of which only 14 required non-invasive oxygen support. Patients underwent chest CT, respiratory function assessment tests, and SARS-CoV-2 IgG antibody titer testing. The present study also used spirometer testing and lung diffusion capacity testing. In the results, no patient had respiratory disease before Covid-19. Three months after discharge, 14.55% experienced effort dyspnea, and 16.36% experienced weakness. Also, in 70.91%, imaging changes in the patients’ lungs were still evident, while 25.45% had some respiratory dysfunction. The dysfunctions were related to TLC in 7.4%, FEV1 in 10.91%, DLCO in 16.36%, and minor airway obstruction in 12.73%. However, the course of respiratory function beyond three months was not reported.

### Hassaan Ahmed et al. (2020)^9^

This meta-analysis aimed to determine long-term clinical outcomes in survivors of severe acute respiratory syndrome (SARS) and Middle East respiratory syndrome (MERS) coronavirus infections after hospitalization or intensive care unit admission. 28 Studies were graded using the Oxford Centre for Evidence-Based Medicine 2009 Level of Evidence Tool. Meta-analysis was used to derive pooled estimates for the prevalence/severity of outcomes up to 6 months after hospital discharge and beyond six months after discharge. Pooled analysis revealed that common complications up to 6 months after discharge were: impaired diffusing capacity for carbon monoxide (prevalence 27%, 95% confidence interval (CI) 15–45%); and reduced exercise capacity (mean 6-min walking distance 461 m, CI 450–473 m).

### Brigitta Fazzini, et al.(2022)^10^

This systematic review and meta-analysis evaluated health-related quality of life (HRQoL), and physical and psychological impairments in ARDS survivors from 3 months to 5-year follow-up after ICU discharge. A Systematic search was done of PubMed, AMED, BNI, and CINAHL databases from January 2000 to date. (n=9992). Regarding the results, Classical ARDS represented 71%, SARS-CoV2 ARDS 27%, and H1N1 ARDS 2%) of the studies. Studies evaluated ARDS survivors for multiple outcomes, including HRQoL (77%), physical function (42%), mental health (35%), pulmonary function (29%), muscle function (19%), cognitive function (12%), return to work (25%), disability (8%), and pain (4%) the 36-Item Short Form Health Survey (SF-36) physical component summary score mean (95% confidence interval [CI]) was 46 (41-50) at 3 months, 39 (36-41) at 6 months, and 40 (38-43) at 12 months. The predictive distance walked in 6 min was 57% (45-69), 63% (56-69), and 66% (62-70) at 3, 6, and 12 months, respectively. Classical ARDS and SARS-CoV-2 ARDS showed no difference in HRQoL and physical function.

**Table 1.** Summary of the studies in the systematic review

| Study | Country and Year of Publication | Type of Study | Aim and Methods | Results |
| --- | --- | --- | --- | --- |
| R Torres-Castro et al.<sup>7</sup> | Spain, 2020 | Systematic review and meta-analysis | To determine the prevalence of restrictive pattern, obstructive pattern and altered diffusion in patients post-COVID-19 infection and to describe the different evaluations of respiratory function used with these patients. A systematic review was conducted in five databases. Studies that used lung function testing to assess post-infection COVID-19 patients were included for review. Two independent reviewers analyzed the studies, extracted the data and assessed the quality of evidence. n= 7 articles reporting on 380 patients | Post-infection COVID-19 patients showed impaired lung function; the most important of the pulmonary function tests affected was the diffusion capacity. In the results, in 40% of the subjects, disturbances in the diffusing capacity of the lungs were observed, in 15% respiratory function limitation was observed and in 7% respiratory function consistent with obstructive disease. |
| Yu-Miao Zhao et al.<sup>8</sup> | Zhengzhou, China, 2020 | Retrospective multicenter cohort study | The relationship between the clinical characteristics and the pulmonary function or CT scores were investigated in ARDS COVID - 19 survivors. n = 55 patients. | SARS-CoV-2 infection related symptoms were detected in 35 of them and different degrees of radiological abnormalities were detected in 39 patients. Lung<br>function abnormalities were detected in 14 patients. Radiological and physiological abnormalities were still found in a considerable proportion of COVID-19 survivors without critical cases 3 months after discharge. |
| Hassaan Ahmed et al.<br>9 | Manchester,<br>2020 | Meta-analysis | To determine long-term clinical outcomes in survivors of severe acute respiratory syndrome (SARS) and Middle East respiratory syndrome (MERS) coronavirus infections after hospitalization or intensive care unit admission. Studies were graded using the Oxford Centre for Evidence-Based Medicine 2009 Level of Evidence Tool. Meta-analysis was used to derive pooled estimates for prevalence/severity of outcomes up to 6 months after hospital discharge, and beyond 6 months after discharge<br>n=28 | Pooled analysis revealed that common complications up to 6 months after discharge were: impaired diffusing capacity for carbon monoxide (prevalence 27%, 95% confidence interval (CI) 15–45%); and reduced exercise capacity (mean 6-min walking distance 461 m, CI 450–473 m) |
| Brigitta Fazzini, et al.<br>10 | London, 2022 | Systematic review and meta-analysis | They evaluated health-related quality of life (HRQoL), and physical and psychological impairments in ARDS survivors from 3 months to 5 yr follow-up after ICU discharge. A Systematic search was done of PubMed, AMED, BNI, and CINAHL databases from January 2000 to date. | The 36-Item Short Form Health Survey (SF-36) physical component summary score mean (95% confidence interval [CI]) was 46 (41-50) at 3 months, 39 (36-41) at 6 months, and 40 (38-43) at |
|  |  |  | n=9992 | 12 months. The predictive distance walked in 6 min was 57% (45-69), 63% (56-69), and 66% (62-70) at 3, 6, and 12 months, respectively. Classical ARDS and SARS-CoV-2 ARDS showed no difference in HRQoL and physical function. |

## Discussion

Experience so far with previous respiratory viruses and ARDS in general shows that patients experience respiratory problems for a long time after hospital discharge. Accordingly, some studies describe prolonged illness from Covid-19 in terms of persistent respiratory symptoms. In addition to the above studies, it is evident that Covid-19 affects respiratory functionality at different levels and severity. According to the study by R Torres-Castro et al., who studied respiratory functionality the first week and then at three (3) months after discharge, using a spirometer, impaired respiratory functionality was noted in terms of lung diffusion capacity in 40% of the participants. In addition, overall respiratory dysfunction was noted in 15% of the participants, while obstructive disorder occurred in 7%. These results are confirmed by Yu-Miao Zhao et al. (2020), a multicentre cohort study, in which it was observed that in a sample of 55 patients three months after discharge, 24.45% experienced some form of respiratory dysfunction while 16.36% experienced impaired CO2 diffusion capacity. An affected diffusivity of CO2 was also noted by Hassaan Ahmed et al. (2020), with an incidence rate of 27%. However, data were collected over a longer period of six (6) months compared to the previous studies, which were conducted over a period of three (3) months. In addition, according to some findings, alterations were observed at the imaging level, i.e., after CT and MRI were performed. In the study by Yu-Miao Zhao et al. (2020), patients underwent chest CT scans where it was found that after a period of three (3) months, there were still imaging findings of Covid-19 disease with concomitant respiratory dysfunction in some of them. However, no similar findings were reported from the other investigations in terms of imaging examinations. Looking further into the respiratory status, the patient’s general physical condition was studied in terms of endurance and functionality, using the 6-min-walking test in combination with spirometry and other similar tests. Subsequently, in the study by Yu-Miao Zhao et al (2020), effort dyspnea was noted at three (3) months from discharge in 14.55% of the participants and weakness in 16.36% of the patients. Accordingly. According to Hassaan Ahmed et al. (2020), at six (6) months from discharge, decreased physical fitness was noted with a mean distance of 6-min walking of 461m. The previous affected reports are also confirmed by Brigitta Fazzini et al. (2022) after conducting a systematic review and meta-analysis of surviving ARDS patients, in whom reduced physical functioning was noted in 42%, disability in 8%, and pain in 4%. In addition, the successful 6-min walking test was performed in 57% at three (3) months, 63% at six (6) months, and 66% at 12 months. Also, at a similar study interval, from the same survey, the following scores were noted in the 36-Item Short Form Health Survey (SF-36): 46 at three months, 39 at six months, and 40 at 12 months; however, these results cannot be compared with other surveys as there was no corresponding evaluation.

## Conclusions

It is a fact that COVID-19 disease, in severe form and following the need for hospitalization due to the development of ARDS, results in an increased likelihood of prolonged occurrence of some symptoms of impaired respiratory function. Impaired CO2 diffusion is observed in most studies, as well as impaired respiratory function in terms of fitness and reduced endurance. Furthermore, the prolonged imaging findings possibly combine with the above research data without a long-term and more massive follow-up. In addition, lower-intensity physical symptoms such as effort dyspnea, fatigue, and pain are observed in the post-hospitalization period. The evolution of symptoms and the general respiratory function of the patient continues to be a major research topic, especially over a five-year period, in order to draw more valid results and conclusions.

## Data Availability

All data produced in the present study are available upon reasonable request to the authors

